# Costs and cost-effectiveness of nurse staffing in acute hospital: a systematic review

**DOI:** 10.1101/2023.02.03.23285139

**Authors:** Peter Griffiths, Christina Saville, Jane Ball, Chiara Dall’Ora, Paul Meredith, Lesley Turner, Jeremy Jones

## Abstract

**Background:** Extensive research shows associations between increased nurse staffing, skill mix and patient outcomes. However, showing that improved staffing levels are linked to improved outcomes is not sufficient to provide a case for increasing them. This review of economic studies in acute hospitals aims to identify costs and consequences associated with different nurse staffing configurations in hospitals.

**Methods:** We included economic studies exploring the effect of variation in nurse staffing. We searched PubMed, CINAHL, Embase Econlit, Cochrane library, DARE, NHS EED and the INAHTA website. Risk of bias was assessed using a framework based on the NICE guidance for public health reviews and Henrikson’s framework for economic evaluations. Inclusion, data extraction and critical appraisal were undertaken by pairs of reviewers with disagreements resolved by a third. Results were synthesised using a hierarchical matrix to summarise findings of economic evaluations

**Results:** We found 22 observational studies conducted in the USA (15), Australia, Belgium, China, South Korea and the United Kingdom (3). Fourteen had high risk of bias and eight moderate. Six studies clearly favoured increased registered nurse staffing with improved outcomes and reduced or unchanged net costs, but most showed increased costs and outcomes. Studies undertaken outside the USA showed that increased nurse staffing was likely to be cost-effective at a per capita GDP threshold or lower. Three studies of skill mix favoured increased registered nurse skill mix, with reduced net cost and similar or improved outcomes.

**Discussion:** Despite the risk of bias, evidence is largely consistent and strongly favours increased skill mix. Evidence for the cost-effectiveness of increases in registered nurse staffing is less certain although it is likely to be cost-effective. In an era of registered nurse scarcity, the results strongly favour investment in registered nurse supply as opposed to using lesser qualified staff as substitutes.

**What is already known?:**

- Higher registered nurse staffing levels and skill mix in acute hospitals are associated with improved care quality and patient outcomes, most notably reduced risk of death
- Previous reviews of evidence strongly support a causal interpretation, but reviews of economic evidence have failed to reach firm conclusions.

**What is study adds:**

- Despite limitations in the evidence, results favour a richer registered nurse skill mix in the nursing workforce as an economically dominant strategy and increases in registered nurse staffing could be cost effective at a low threshold

**How this study might affect research practice or policy:**

- Workforce substitution is unlikely to provide a cost-effective solution to registered nurse shortages.
- More incremental cost effectiveness studies are required to guide decision making in context as specific results do not generalise

**CREDiT (author contributions):** Jane Ball, Chiara Dall’Ora, Peter Griffiths, Jeremy Jones, Paul Meredith acquired funding for the study Peter Griffiths, Jeremy Jones, Christina Saville designed the review and wrote the protocol

Christina Saville undertook the searches

Jane Ball, Jeremy Jones, Paul Meredith, Lesley Turner, Christina Saville undertook screening and application of the inclusion criteria

Jane Ball, Chiara Dall’Ora, Peter Griffiths, Jeremy Jones, Paul Meredith, Lesley Turner, Christina Saville undertook data extraction and critical appraisal of included studies

Christina Saville and Peter Griffiths prepared the first draft of the paper

Peter Griffiths undertook data synthesis and drafted results

Jane Ball, Chiara Dall’Ora, Peter Griffiths, Jeremy Jones, Paul Meredith, Lesley Turner, Christina Saville reviewed the first draft and provided critical comment and contribution to the discussion / suggested revisions.

All reviewed the final draft

**Registration:** PROSPERO (CRD42021281202).

## 1 Introduction

Many countries face significant shortages of registered nurse supply, motivating calls for further investment in nurse training or the search for alternative ways of staffing wards, including the creation of new cadres of nursing staff with lower levels of qualifications and increased use of unregistered support staff.^1 2^ There is substantial evidence demonstrating that patients in hospitals with more registered nurses experience higher quality care and have lower risk of complications and death. However, the value of this information for guiding policy and operational decisions has been questioned. ^3^ In the face of competing demands for scarce financial and labour resources, economic evaluations are required to inform decision-making.

Several reviews have summarised evidence linking higher nurse staffing and skill-mix to improved patient outcomes and quality, finding hundreds of studies from around the world.^4-7^ The evidence is almost exclusively observational but there is a clear consensus that the associations observed are at least partly causal. Findings from hospital level cross-sectional studies are increasingly supported by longitudinal patient level studies showing effects from exposure to low staffing.^4^ Effect sizes are, however, typically small. Exposure to low staffing (variously defined) on general medical / surgical units typically increases the hazard of death by 2-3%.^4^

With nurse staffing comprising a large proportion of the pay bill for hospitals the cost-effectiveness of nurse staffing improvements relative to other potential investments should not be assumed, although increased staffing may reduce costs associated with treating complications.^8^ Existing reviews have found a relatively small number of economic studies, which were hard to synthesise due to the differing methods and measures used.^3 9^ The underlying observational studies used to estimate effectiveness, and the limited economic perspective taken, have been noted as key limitations. Most effectiveness estimates come from cross-sectional studies and few studies consider costs beyond the immediate hospital stay. ^9^ These reviews are now dated and there is significant new evidence. In this paper we aim to provide an up-to-date review of economic studies of nurse staffing and skill mix in acute hospitals to identify the costs and consequences associated with different nurse staffing configurations in hospitals.

## 2 Methods

### 2.1 Study eligibility and search strategy

We included economic studies exploring the effect of variation in nurse staffing in acute hospital inpatient settings. We included studies that measured variation in staffing level (e.g. nurse-patient ratio, nurse hours per patient day), understaffing (e.g. nurse staffing below specified threshold) or skill mix (e.g. RN/all nursing staff ratio). Nurse staffing included any or all staff working as part of a nursing care delivery team in an inpatient unit (including registered nurses [RN] and nursing aides / assistants). For studies in maternity settings, such as post-natal wards we also included registered midwives. We excluded studies exclusively conducted in psychiatric/mental health care, community or long-term care and emergency departments.

We included cost minimisation, cost-benefit, cost consequences, cost-effectiveness and cost utility studies conducted as part of prospective intervention studies (including randomised and quasi-experimental designs), observational studies and secondary modelling studies.

Given the variety of economic evaluation approaches, there were a range of possibilities for how costs and consequences might be evaluated, and we did not limit the study selection in this regard although studies had to provide a direct monetary cost (as opposed to un-costed measure of resource use). Cost-minimisation studies that simply compared staff costs were not considered although studies that compared net-costs of different staffing strategies were.

We searched PubMed, CINAHL, Embase, Econlit, Cochrane library (CDSR, CENTRAL, Protocols), Database of Abstracts of Reviews of Effectiveness (DARE), NHS EED) and the INAHTA website (HTA) up to October 2022. Search terms are provided in Supplementary Table 7. We included all eleven studies from two existing systematic reviews ^9 10^ and considered additional relevant texts found in authors’ existing reference libraries and in the reference lists of seminal papers. We included peer-reviewed journal articles, theses and conference proceedings published in English and we found no titles / abstracts of non-English papers that could have been eligible. The review was registered on PROSPERO (reference: CRD42021281202).

### 2.2 Data extraction

The initial search, deduplication and title/abstract screening were conducted by one reviewer. Two reviewers reviewed full texts independently, and disagreements were resolved by discussion with the entire review team. Data extraction was undertaken by one reviewer and checked by a second. From each study we extracted author(s), year, country and setting, study design, sample size, staff group(s), source of staffing data, measure of staffing levels/skill mix, natural variation/ planned change, level of aggregation at which staffing measured and analysed, economic perspective, time horizon over which the consequences of nurse staffing variation are evaluated, costs and relevant outcomes. We extracted both costs and consequences when they were reported in a disaggregated fashion. We extracted mortality-based cost-effectiveness estimates or calculated incremental cost per life saved from the data reported. We translated these estimates into US$ and a common year for ease of comparison, first converting to 2021 costs using country specific inflation, then converting to equivalent costs in US$ using OECD purchasing power parity tables. ^11^

### 2.3 Quality appraisal

We assessed the risk of bias in the underlying studies using a framework based on that used for the development of NICE public health guidance. ^10 12^ Because most studies used routine administrative data for resource use and clinical outcomes, we focussed our assessment on items related to the underlying study design, sample size, representativeness of patients hospitals and staff samples (in relation to the target populations for inference from the study) and control for confounding (see supplementary Appendix 1). We did not calculate summary scores, but we gave overall assessments based on the lowest scoring items for the domains of internal and external validity and across all items (internal and external validity combined). Risk of bias assessments were undertaken by two reviewers with disagreements resolved with reference to a third.

We additionally used Henrikson’s framework, which brings together common domains from three economic reporting checklists.^13^ Because there was considerable overlap with the domains covered when assessing the underlying studies, and almost all studies derived data from administrative systems, we focussed particularly on assessing the comprehensiveness of cost / resource use, including the cost perspective, time horizon, and tests for sensitivity to key assumptions about costs linked to precision and underlying bias of staffing outcome association estimate. We classified costs included as direct staff costs, general consequential (due to changes in length of stay), additional treatment costs, post discharge care costs and societal costs. We gave a summary of the relative comprehensiveness of costs considered by summing the areas of cost covered. We classified the strength of the approach to economic analysis for decision making, ranging from lowest (cost minimisation) low (cost consequences) moderate (cost-effectiveness) to high (cost utility / cost benefit).

The diversity of the evidence made a formal assessment of publication bias unfeasible, but the issue and likely biases were addressed in narrative discussion. Similarly, we did not formally assess overall of strength of evidence / recommendations using GRADE ^14^ but used it to shape our discussion.

### 2.4 Synthesis

We considered statistical meta-analysis but heterogeneity in terms of interventions, range of costs considered and health economies among the studies we found led us to focus on qualitative reporting for synthesis. We performed a narrative synthesis, considering patterns of results. To support this we developed graphical displays based on a hierarchical matrix to summarise findings of economic evaluations, as described by Nixon and colleagues. ^15^ Constellations of results (increases / decreases in costs, improvements / decline in outcomes) are organised by the economic decisions that arise. In a classic health economic decision-making framework, where costs are increased and health outcomes are not improved, or if costs are unchanged and outcomes are worsened then an intervention should be rejected on economic grounds. Conversely if outcomes are improved and costs are not increased, the intervention should be accepted. Other results, typically where improved outcomes are associated with increased costs, an incremental cost-effectiveness analysis is required to inform decision making to determine whether an intervention should be accepted.

As there is no universally accepted threshold to establish cost-effectiveness, we tabulated mortality based incremental cost-effectiveness estimates against the countries per capita gross domestic product (GDP). A cost of 1 X the per capita GDP per quality adjusted life year is sometimes used as a threshold for defining cost-effectiveness, although many consider that this may be excessive.^16 17^ Therefore we used it as a reference point, providing an upper bound for potential cost-effectiveness. If increases in staffing yield a cost per quality adjusted life year more than the per-capita GDP, it is unlikely to be considered cost-effective by any criteria.

## 3 Results

We identified 6783 studies from database searches and fourteen from other sources. 68 were retained after title and abstract screening and we included 23 papers reporting on 22 studies. See Figure 1 for PRISMA flow chart and Table 1 for details of the studies.

**Table 1.** Characteristics of included studies

| Paper | Country | Patient group | Study design * | Source of variation | Level of aggregation for staffing | Hospitals | Patients | Economic analysis |
| --- | --- | --- | --- | --- | --- | --- | --- | --- |
| Behner et al. 1990 <sup>18</sup> | USA | Back and neck procedures | Retrospective observational study | Natural variation | Patient stay | 1 | 132 | Cost - consequences (disaggregated) & net cost of avoiding low (20% below standard) staffing |
| Clark et al. 2014 <sup>19</sup> | USA | Maternity - induction of labour | Retrospective cross-sectional observational study | Natural variation | Hospital | 110 | 101377 | Cost - consequences (disaggregated) of providing universal 1:1 midwifery care |
| Cookson et al. 2014 <sup>20</sup> | UK | Maternity | Retrospective cross-sectional observational study | Natural variation | Hospital | 157 | 5,753,551 | Cost-effectiveness (1 additional midwife per 100 deliveries) |
| Dall et al. 2009 <sup>21</sup> | USA | General med /surg | Secondary modelling (data from cross-sectional studies)<sup>a</sup> | Simulated change | Hospital | 610 | 5,400,000 | Cost - (monetary) benefits (disaggregated) and consequences of increased RN staffing |
| Griffiths et al. 2018 <sup>22</sup> | UK | General med /surg | Retrospective longitudinal observational | Natural variation | Patient day | 1 | 138133 | Cost-effectiveness per additional RN hour per patient day and increasing skill mix to establishment |
| Griffiths et al. 2020 <sup>23 24</sup> | UK | General med /surg | Simulation model (parameter data from retrospective longitudinal study)<sup>b</sup> | Simulated change | Shift | 4 | NA | Cost consequences (low staffing) and effects of different baseline staffing policies |
| Kim et al. 2016 <sup>25</sup> | South Korea | Hip & knee surgery | Retrospective cross-sectional observational | Natural variation | Hospital | 222 | 22289 | Care Cost (charges) consequences (disaggregated) of different patient:RN ratios in hospitals |
| Lasater et al. 2021a <sup>26</sup> | USA | General med /surg (select) | Retrospective cross-sectional observational | Natural variation | Hospital | 116 | 417861 | Cost - consequence (disaggregated) of changed patient:RN ratio |
| Lasater et al. 2021b <sup>27</sup> | USA | General surg (select) | Matched cohort / Retrospective cross-sectional observational | Natural variation | Hospital | 306 | 125430 | Cost - effectiveness of composite nursing resource (staffing, skill mix, BSN mix and nurse reported work environment) |
| Lasater et al. 2021c <sup>28</sup> | USA | General med (select) | Matched cohort / Retrospective cross-sectional observational | Natural variation | Hospital | 306 | 148090 | Cost - effectiveness of composite nursing resource (staffing, skill mix, BSN mix and nurse reported work environment) |
| Li et al. 2011 <sup>29</sup> | USA | General med /surg | Retrospective cross-sectional observational | Natural variation | Unit | 125 | 110646 | Costs of additional nursing hour and increased RN skill mix |
| Li et al. 2016 <sup>30</sup> | USA | Cardiac surgery | Propensity matched cohort / Retrospective cross-sectional observational | Natural variation | Hospital | 1887 | 439365 | Cost - consequence (disaggregated) of hospital above median staffing (RHPPD≥7.07) vs below (HPPD < 7.07) |
| Martsof et al. 2014 <sup>31</sup> | USA | General med /surg | Matched cohort / Retrospective cross-sectional observational | Natural variation | Hospital | 421 | 18474860 | Cost - consequence (disaggregated) of additional nurse (RN/LPN) per 1000 admissions and higher RN / Licensed skill mix |
| Needleman et al. 2006 <sup>8</sup> | USA | General med /surg | Secondary modelling (data from cross-sectional studies)<sup>a</sup> | Simulated change | Hospital | 799 | 5075969 | Cost - consequences (disaggregated) of different staffing levels / configurations (HPPD) |

*Economic evaluations of nurse staffing*
| Paper | Country | Patient group | Study design * | Source of variation | Level of aggregation for staffing | Hospitals | Patients | Economic analysis |
| --- | --- | --- | --- | --- | --- | --- | --- | --- |
| Pang et al. 2019 <sup>32</sup> | China | Neurology / neurosurgery | Prospective cross-sectional observational | Natural variation | Hospital | 1 | 5091 | Cost-consequence (disaggregated) of care in 6 wards with different proportion of RNs. |
| Ross et al. 2021 <sup>33</sup> | USA | Pulmonary lobectomy | Retrospective cross-sectional observational study | Natural variation | Hospital | NA | 16944 | Cost-consequence (disaggregated) of different staffing levels (RN FTEs per 1000 patient days) |
| Rothberg et al. 2005 <sup>34</sup> | USA | General med /surg | Secondary modelling* (data from cross-sectional studies)<sup>c</sup> | Natural variation | Hospital | 799 | 5075969 | Cost-effectiveness per unit reduction in patient to nurse ratio |
| Shamliyan et al. 2009 <sup>35</sup> | USA | General med /surg | Secondary modelling* (data from cross-sectional studies)<sup>d</sup> | Natural variation | Hospital | NA | NA | Net benefit and Cost-Benefit arising from avoided deaths (and adverse events - not reported) corresponding to a 1 FTE RN per 1000 patients increase |
| Twigg et al. 2013 <sup>36</sup> | Australia | General med /surg | Retrospective observational study | Implementation of new staffing levels | Hospital | 3 | 214261 | Cost-effectiveness of implementing a NHPP method to guide nurse staffing |
| Van den Heede et al. 2010 <sup>37</sup> | Belgium | Cardiac surgery | Secondary modelling* (data from cross-sectional studies) | Simulated change | Ward | 28 | 9054 | Cost-effectiveness of increasing nurse staffing to the 75th centile |
| Weiss et al. 2011 <sup>38</sup> | USA | General med /surg | Prospective cross-sectional observational | Natural variation | Ward | 4 | 1892 | Cost consequences (disaggregated) of increasing non overtime RN staffing |
| Yakusheva et al. 2014 <sup>39</sup> | USA | General med /surg | Retrospective observational study | Simulated change | Patient stay | 1 | 8526 | Cost consequences (disaggregated) of increasing % BSN qualified RN staffing |

- study providing effectiveness estimates a. Needleman 2001, 2002 <sup>40 41</sup> b. Griffiths 2018 <sup>22</sup> c Aiken et al 2002 <sup>42</sup> d Kane et al 2007 <sup>5</sup>.

**Figure 1.**
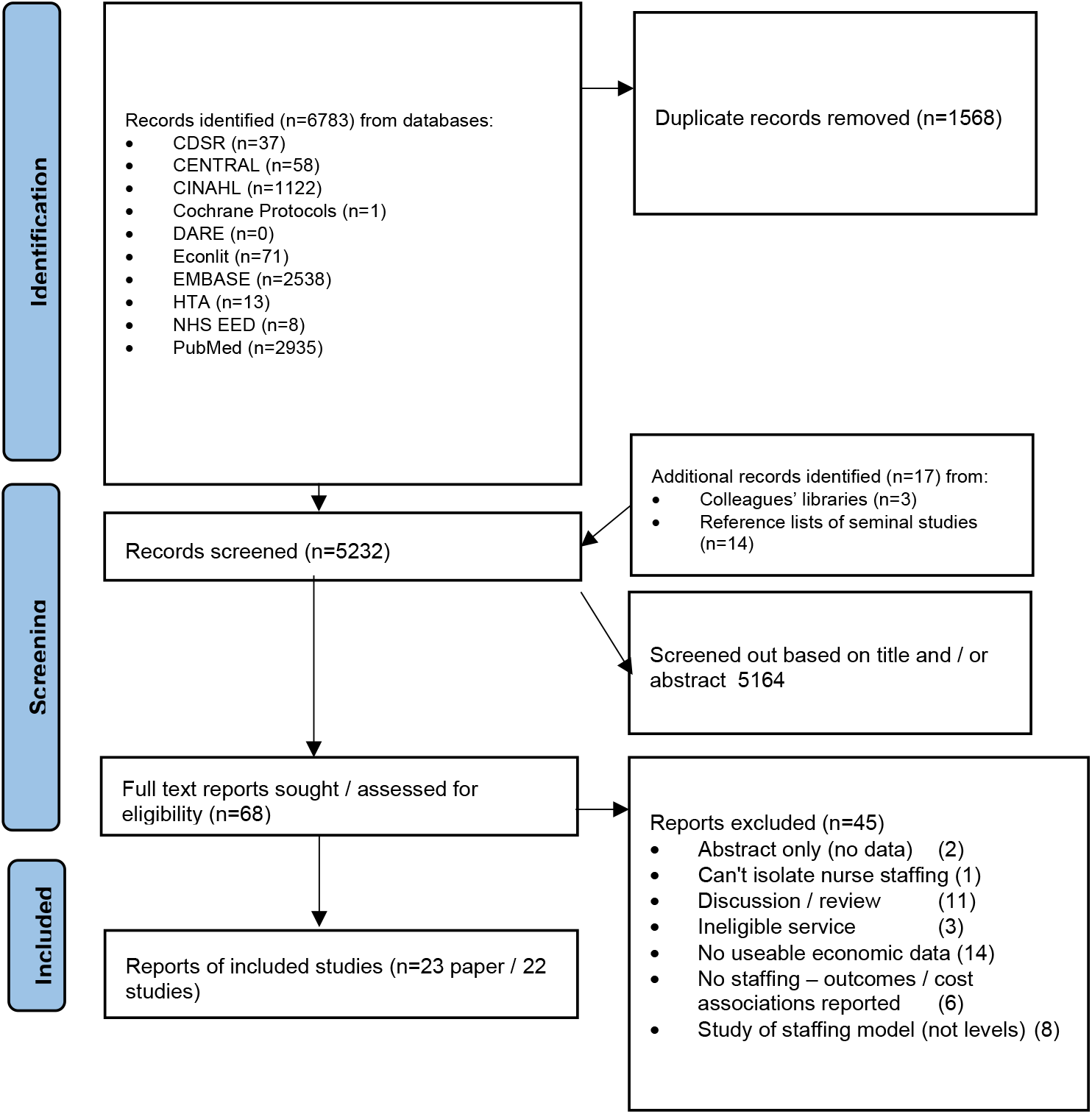
PRISMA flow diagram

### 3.1 Study characteristics

Most studies (17) addressed staffing on general medical and / or surgical units while the rest addressed specific surgical specialties or procedures ^18 25 29 32 33^ or maternity care.^19 20^ Publications were from 1990 to 2021 although data in some studies were considerably earlier than publication year. Most studies (15/22) were conducted in the USA ^8 18 19 21 26-31 33-35 38 39^, three in the UK ^20 22-24^, and one each in Australia ^36^, Belgium^37^, China ^32^ and South Korea ^25^. In total, data came from 5900 hospitals and over 41 million patients. See Table 1

All studies were observational or sourced parameters and data for modelling from observational studies. Most studies used estimates of effects based on natural variation in registered nurse or midwife staffing expressed as a staff to patient ratio (or vice versa), using staffing outcome associations to model the effect of various changes in staffing levels. Of these, three used parameters from natural variation to model the effects of planned change across health systems ^8 21 37^, (typically increasing staffing to the 75^th^ centile). One study used a mathematical simulation model to explore the effects of different approaches to determining staffing levels on achieved staffing. ^23 24^ In one study the observed variation in staffing arose from implementing a method to determine staff requirements which led to increased staffing levels. ^36^

### 3.2 Study quality and risk of bias

Eight studies were rated as moderate risk of bias with the remaining rated as having a high risk of bias (see Table 2 & supplementary Table 3). Because of large sample sizes and risk adjustment, most studies were assessed as low risk of bias related to power and control of confounders but the intrinsic design limitations of cross-sectional studies meant that only two studies (reported in three papers) were rated as strong for internal validity, ^22-24^ with a further six rated as moderate.^27 28 30 35 36 39^ Most studies (18) used estimates for the effect of nurse staffing that were cross-sectional in the sense that staffing levels are aggregated over a large unit (typically a hospital) over time (typically a year) and linked with outcomes of patients admitted over that period. Of these, four use potentially stronger matched cohort designs. ^27 28 30 31^ Four studies provide (or use) estimates of staffing / outcome associations that directly link patients to staffing at a day or shift level (or equivalent) ^18 23 24 39^ and one compared outcomes before and after a planned change in staffing.^36^ Despite the preponderance of large multi-hospital studies, only seven studies were rated as potentially strong for external validity ^8 20 21 29 31 35 37^ with a number of large studies down-graded to moderate risk of bias because there was a mismatch between the patient sub-group providing outcomes and the patient population served by the staff included.

**Table 2.** Assessment of economic study quality

| Study | Economic perspective | Discounting / time horizon? | Staff cost | Consequential | Treatment cost | Post-discharge cost | Societal costs | Sensitivity analysis | Range of costs<sup>a</sup> | Level of economic analysis<sup>b</sup> | Risk of bias in underlying study |
| --- | --- | --- | --- | --- | --- | --- | --- | --- | --- | --- | --- |
| Behner et al. 1990 <sup>18</sup> | hospital | n / i | Salary | hospital stay | Adverse events |  |  | no | Moderate | Moderate | High |
| Clark et al. 2014 <sup>19</sup> | hospital | n / i | Salary |  |  |  |  | no | Limited | Moderate | High |
| Cookson et al. 2014 <sup>20</sup> | hospital | n / i | Employment |  |  |  |  | yes | Limited | Moderate | High |
| Dall et al. 2009 <sup>21</sup> | societal | y / life | Employment | hospital stay | AE | follow up care and treatment | productive value | no | Extensive | Moderate | High |
| Griffiths et al. 2018 <sup>22</sup> | hospital | n / i | Employment | hospital stay |  |  |  | yes | Moderate | Moderate | Moderate |
| Griffiths et al. 2020 <sup>23 24</sup> | hospital | n / i | Employment | hospital stay |  |  |  | yes | Moderate | Moderate | Moderate |
| Kim et al. 2016 <sup>25</sup> | hospital | n / i | Charges | hospital stay |  |  |  | yes | Moderate | Moderate | High |
| Lasater et al. 2021a <sup>26</sup> | hospital / patient | n / i | Hospital costs |  |  | readmission |  | no | Moderate | Moderate | High |
| Lasater et al. 2021b <sup>27</sup> | hospital / patient | n / i | Employment | hospital stay |  | readmission |  | no | Moderate | Moderate | Moderate |
| Lasater et al. 2021c <sup>28</sup> | hospital / patient | n / i | Employment | hospital stay |  | readmission |  | yes | Moderate | Moderate | Moderate |
| Li et al. 2011 <sup>29</sup> | hospital | n / i | Hospital costs |  |  |  |  | yes | Limited | Low | High |
| Li et al. 2016 <sup>30</sup> | hospital | n / i | Hospital costs | hospital stay | AE |  |  | yes | Moderate | Moderate | Moderate |
| Martsof et al. 2014 <sup>31</sup> | hospital | n / i | Hospital costs | hospital stay | AE |  |  | yes | Moderate | Moderate | High |
| Needleman et al. 2006 <sup>8</sup> | hospital | n / i | Salary | hospital stay | AE |  |  | no | Moderate | Moderate | High |
| Pang et al. 2019 <sup>32</sup> | hospital | n / i | Employment |  |  |  |  | no | Limited | Moderate | High |
| Ross et al. 2021 <sup>33</sup> | hospital | n / i | Hospital costs | hospital stay |  |  |  | no | Moderate | Moderate | High |
| Rothberg et al. 2005 <sup>34</sup> | hospital | n / i | Salary | hospital stay |  |  |  | yes | Moderate | Moderate | High |
| Shamliyan et al. 2009 <sup>35</sup> | societal | y / life | Employment | hospital stay | AE |  | future earnings | yes | Extensive | High | Moderate |
| Twigg et al. 2013 <sup>36</sup> | hospital | y / life | Employment |  | AE |  |  | yes | Moderate | Moderate | Moderate |
| Van den Heede et al. 2010 <sup>37</sup> | hospital | y / life | Salary |  |  |  |  | yes | Limited | Moderate | High |
| Weiss et al. 2011 <sup>38</sup> | hospital / payer | n / i | Employment |  |  | readmission |  | no | Moderate | Moderate | High |
| Yakusheva et al. 2014 <sup>39</sup> | hospital | n / i | Salary |  |  | readmission |  | yes | Moderate | Moderate | Moderate |

<sup>a</sup> Limited – 1 cost domain – Moderate 2 or 3 cost domains – Extensive 4 or 5
<sup>b</sup> Low – cost only study, Moderate cost consequences or cost effectiveness, High cost utility or cost benefit

The economic analysis in most studies was a disaggregated cost consequences analysis with a range of consequences reported – typically restricted to some or all of mortality rates, adverse incidents, length of stay and readmissions (see Table 2). The economic perspective was that of the hospital in most studies and in five studies only staffing costs were considered. ^19 20 29 32 37^ The remainder considered at least some consequential costs ranging from costs of extended stays, treatments of adverse events, readmissions and, in two cases, societal costs in terms of lost earning or productive capacity. ^21 35^ While most studies took an immediate perspective on both outcomes and cost, two took a lifetime perspective on outcomes ^36 37^, estimating life expectancies, while two ^21 35^ considered lifetime future earnings / productivity. We were able to extract or calculate a cost-effectiveness ratio related to death as an outcome from 8 studies ^8 21-23 27 28 36 37^ and one study provided cost benefit analysis in terms of a ratio of staff costs to financial benefits arising from care cost savings and future productivity. ^35^ While many studies undertook some form of sensitivity analysis, estimates of economic parameters did not reflect underlying uncertainty (e.g. 95% confidence intervals).

### 3.3 Costs and cost-effectiveness

For details of the main economic results see Table 3. In all cases simple increases in staff led to increased staffing costs, as did increases in skill mix. Sixteen studies provided estimates of net costs associated with staffing increases, taking into account other costs / savings that might result from staff changes. Of these, five found that increases in registered nurse staffing in general medical / surgical or other surgical specialities led to reduced costs overall. ^18 25 26 35 38^ All but one of these studies were rated as high risk of bias. The other study, with moderate risk of bias, found that economic benefits to society, including losses to productivity avoided, exceeded costs with a benefit to cost ratios for each additional RN between 1.27 and 2.51. One additional RN per ‘000 surgical patients in US hospitals cost $923,832 but yielded a benefit of $1,646,190. For medical and intensive care costs of an additional RN per ‘000 patients ($982,800 / $589,680) were also less than benefits ($1,244,061 /$1,479,933). ^35^ Two studies in US general medical / surgical patients found no net cost difference from staff increases ^28 31^ while the remaining nine found net cost increases ^8 21-24 27 29 30 33 34^. In all but one study, there was evidence of improved health outcome associated with increased staffing. Clark et al. 2014 found increased costs but no evidence of reduced complications from increased nurse staffing to achieve 1 to 1 staffing during induction of labour in US maternity settings. ^19^

**Table 3.**
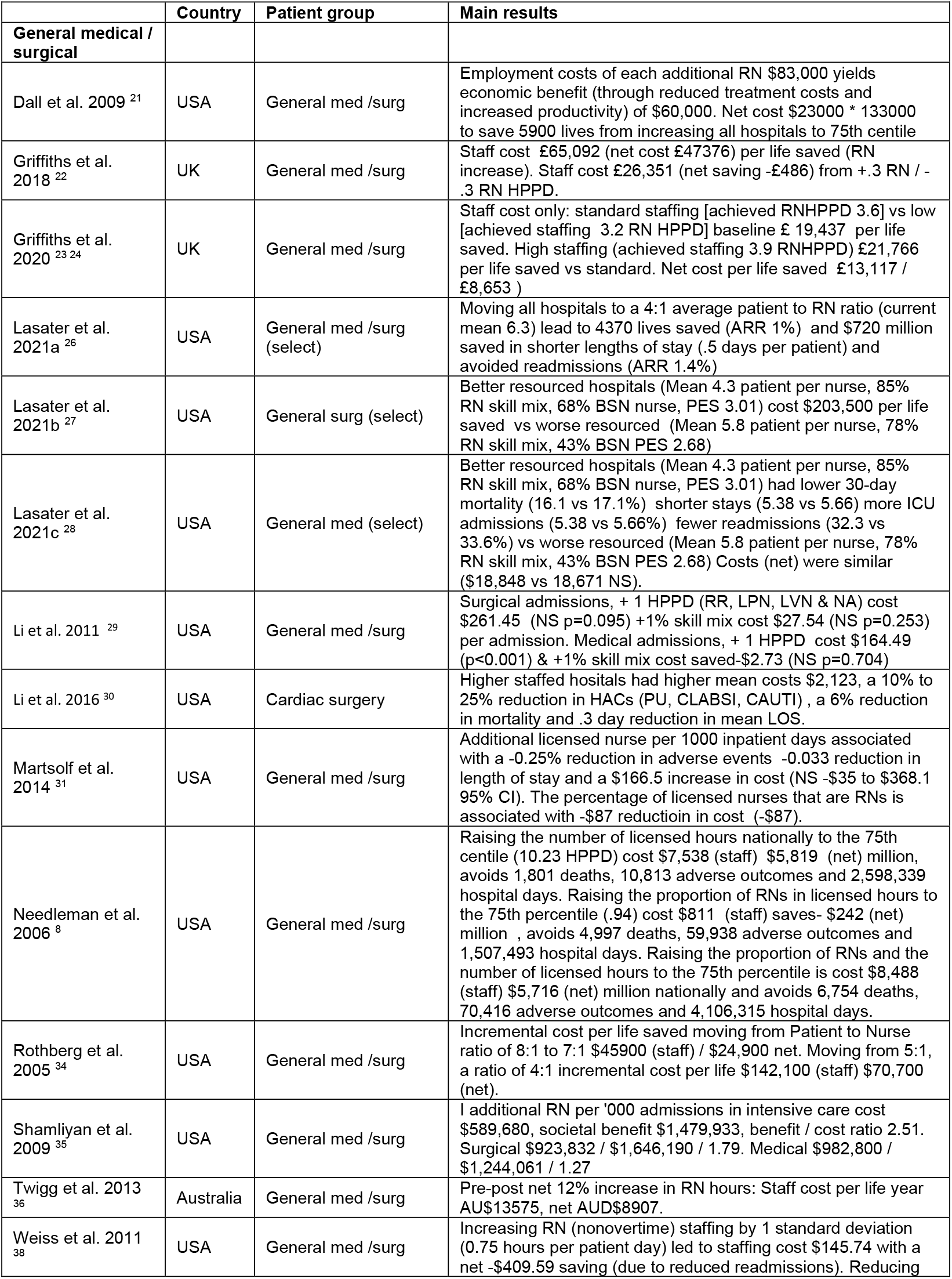
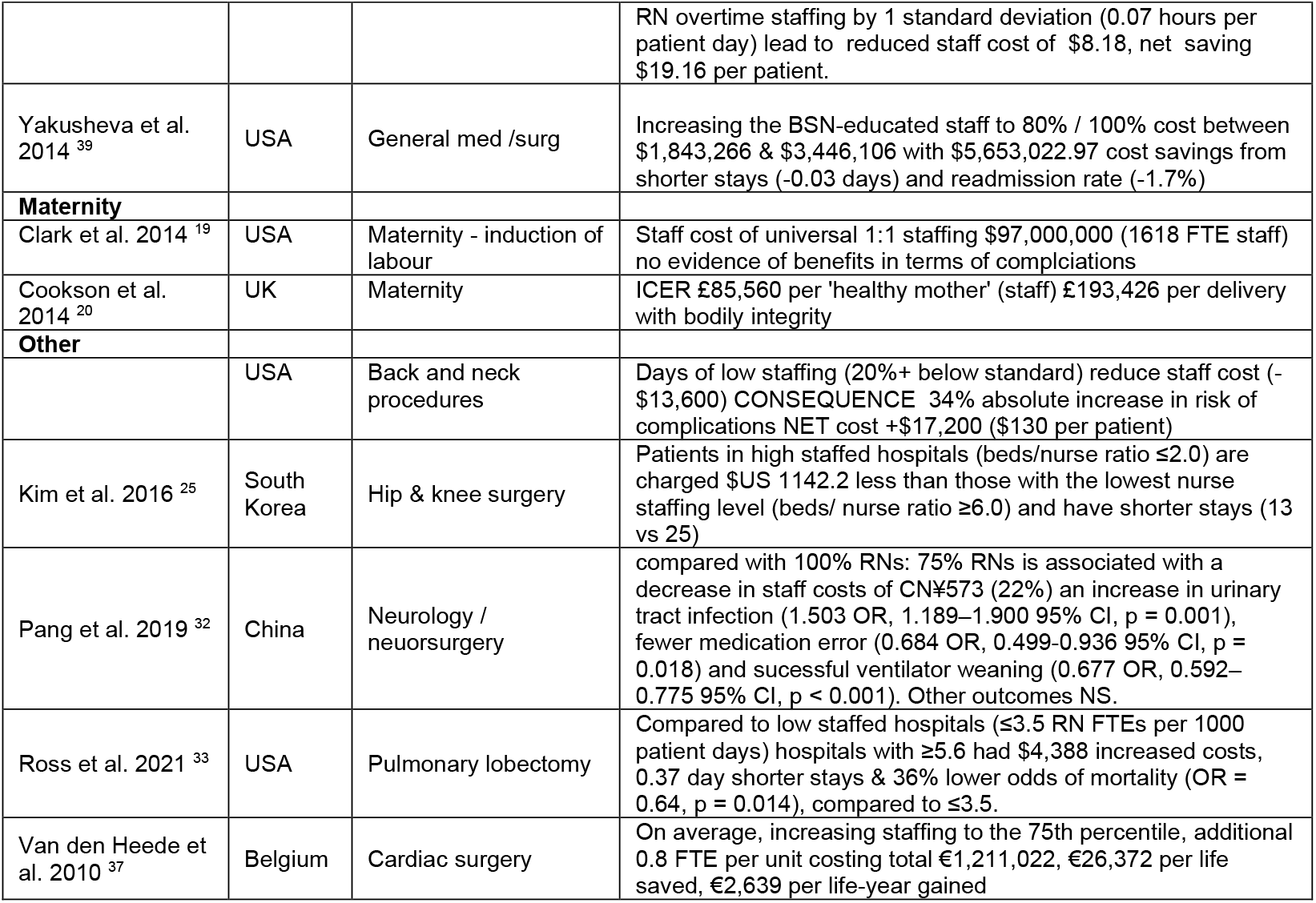
Summary of results

For change of skill mix, all four studies that considered net costs found that a skill mix that was richer in registered nurses was associated with reduced net costs overall. ^8 22 29 31^ Four studies found improved health outcomes from increasing the proportion of RNs in the nursing team in medical / surgical settings (three general and one neurology / neurosurgery) while fifth found that increasing the proportion of bachelors educated RNs was associated with improved outcomes. ^39^ Of these studies two were assessed as moderate risk of bias with one rated as low risk of bias based on internal validity, although both were single site studies. ^22 39^

Figure 2 summarises results in a hierarchical matrix.15 In total six studies provided results that clearly supported increased RN staffing when using net costs, with a combination of no cost change but improved outcomes ^28 31^ or reduced costs and improved outcomes ^18 26 35 38^ in medical and / or surgical wards. Of these, two studies were rated as moderate for the underlying risk of bias. ^28 31^ However, most studies showed both increased costs and improvements in health outcomes, where incremental (cost-effectiveness) analysis is required to inform the economic decision. All three studies that considered net costs supported a decision to increase skill-mix ^8 22 31^, including one study with moderate risk of bias, although if using staff costs alone the results of four studies with improved outcomes and increased costs mean that incremental analysis is required for decision making. For increases in staffing levels, a single study gave results that clearly reject staffing increases, but this used limited cost data and was at high risk of bias.^19^

**Figure 2:**
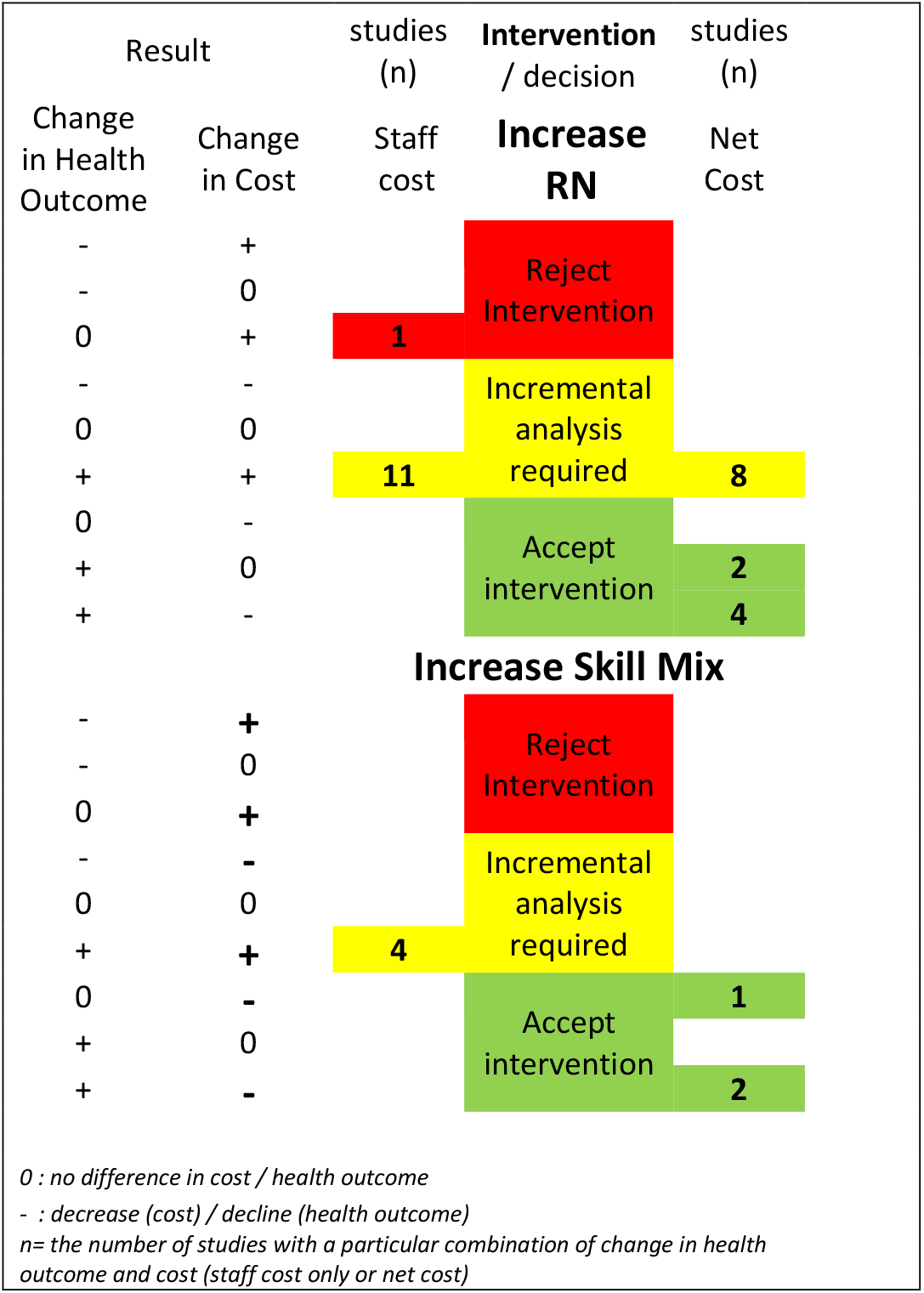
hierarchical matrix to summarise findings & economic conclusions from economic studies of nurse staffing / RN skill mix -increase.

In Table 4 the cost-effectiveness estimates are summarised, alongside the 2021 per capita gross domestic product (GDP) of the country providing the estimate. Twigg et al. 2013 ^36^ & Van den Heede et al. 2010 ^37^ provide cost per life year. In both cases the cost per life year is far below per capita GDP and adjustment for loss of utility (quality) is unlikely to substantively alter the conclusions that the staffing increases (in Australian general medical / surgical units and Belgium cardiac units respectively) are likely to be cost-effective at a GDP based threshold. For four studies in US and UK general medicine / surgery the ratio between per capita GDP / cost per life saved ranged from 0.3 ^28^ to 3.2 ^27^, although both the US studies provided estimates for a ‘combined’ intervention, implying both increased RN staffing and additional changes in skill-mix beyond that which would result from the staff increases. Even the higher end of this range is potentially cost-effective if each ‘life saved’ gains 3.2 quality adjusted years. Other US studies require that each life saved yield more than 12 QALYs ^21^ or, in the case of Needleman et al ^8^ nearly 70 QALYs to achieve the GDP based threshold.

**Table 4.** Costs per life saved from studies of increased staffing

| Paper | Country | Patient group | Main results | Cost per life (*life year) saved (2021 equivalent) | Cost per life (*life year) saved (2021 US\$ / purchasing power parity equivalent. | 2021 per capita GDP (in US\$) |
| --- | --- | --- | --- | --- | --- | --- |
| Dall et al. 2009 <sup>21</sup> | USA | General med /surg | Increasing RN staffing in all hospitals to 75th centile | US\$ 839,930 | \$ 839,930 | \$69,287.5 |
| Griffiths et al. 2018 <sup>22</sup> | UK | General med /surg | Increase of 1 RN Hour per patient day | GB£ 54,009 | \$ 77,957 | \$47,334.4 |
| Griffiths et al. 2020 <sup>23 24</sup> | UK | General med /surg | Standard staffing policy [achieved RNHPPD 3.6] vs low staffing [achieved staffing 3.2 RN HPPD] | US\$ 14,560 | \$ 21,016 | \$47,334.4 |
| Lasater et al. 2021b <sup>27</sup> | USA | General surg (select) | Better resourced hospitals (Mean 4.3 patient per nurse, 85% RN skill mix, 68% BSN nurse) vs worse resourced (Mean 5.8 patient per nurse, 78% RN skill mix) | US\$ 221,815 | \$ 221,815 | \$69,287.5 |
| Lasater et al. 2021c <sup>28</sup> | USA | General med (select) | Better resourced hospitals (Mean 4.3 patient per nurse, 85% RN skill mix, 68% BSN nurse) vs worse resourced (Mean 5.8 patient per nurse, 78% RN skill mix, 43% BSN) | US\$ 18,127 | \$ 18,127 | \$69,287.5 |
| Needleman et al. 2006 <sup>8</sup> | USA | General med /surg | Raising the number of licensed hours nationally to the 75th centile | US\$ 4,840,377 | \$ 4,840,377 | \$69,287.5 |
| Twigg et al. 2013 <sup>36</sup> | Australia | General med /surg | Implementation of RN hours per patient day staffing model - net 12% increase in RN hours. | AU\$12,114* | \$ 8,418 * | \$ 59,934.1 |
| Van den Heede et al. 2010 <sup>37</sup> | Belgium | Cardiac surgery | Increasing staffing to the 75th percentile (additional 0.8 FTE per unit) | € 3,510* | \$ 4,726* | \$ 51,767.8 |

## 4 Discussion

We have identified economic evaluations of change in the size and / or composition in nursing midwifery staff in hospitals and have found considerably more evidence than considered in previous reviews, which have proved inconclusive. The evidence is extensive with twenty-two studies using data from many millions of patients over many countries. The largest body of evidence relates to registered nurse staff levels in adult medical and / or surgical wards with a smaller number of studies addressing skill mix. Most studies found that staffing increases were either dominant or provided results consistent with cost-effectiveness based on a per capita gross domestic threshold. In many cases staffing increases were consistent with cost-effectiveness at a considerably lower threshold. In the majority of studies exploring skill mix, increased skill mix (higher proportion of RNs or increased qualification of RNs) was an economically dominant strategy based on consideration of net costs. However, evidence came from diverse contexts and evaluated a range of different interventions and the quality of the underlying observational studies had, at best, a moderate risk of bias.

Decisions about the cost-effectiveness of nurse staffing increases are contingent and evidence from local contexts is desirable. Nonetheless all evidence from countries other than the USA gave results that are compatible with cost-effectiveness at a per capita GDP per quality adjusted life year-based threshold. In the simplest case, a blanket one hour per patient per day increase in RN staffing in the UK cost $77,957 per life saved (2021 US$ equivalent) ^22^ in the context of a per capita GDP of $47,334, which would be cost effective if each life saved gained 1.6 quality adjusted life years. Discounted quality adjusted life expectancy for an 80-90 year old with comorbidities in the UK is estimated to be over 2 years ^43^, with over 6 years estimated for a population similar to the inpatient population at risk. ^44^ Other cost-effectiveness estimates were based on more complex staffing changes, such as bringing all hospitals up to a defined level of staffing, improved staffing and skill mix combined or changed baseline staff establishments to meet varying need. In general, these results were more favourable to increased staffing, indicating that cost-effectiveness may be enhanced by targeted intervention focussed on areas with greater deficit or guided by validated staffing tools.

However, in a resource constrained system, consideration must be given to the opportunity costs when considering whether or not the health benefits gained are greater than the health that is likely to be lost because resources are not deployed elsewhere.^17^ Substantially lower thresholds have been proposed. In the UK context the National Institute for Health and care Excellence, the body charged with assessing evidence to inform health care provision in the publicly funded health system identified £10,000 per QALY ($15,572 2021 US$ equivalent) as representing ‘exceptional value for money’. In most cases it seems likely that staff increases could be cost-effective at this lower threshold. To this must be added the weight of six studies where staff increases were associated with improved outcomes and reduced net costs, where the decision to increase staffing dominates.

The major exception to a conclusion of likely cost-effectiveness comes from two US studies where costs per life saved from RN staffing increases in general medical surgical units are many multiples of per capita GDP. ^8 21^ Other US studies suggest that even at this level there may be net societal benefit once lost productivity is considered. ^21 35^ While this societal perspective is important, it may have less influence on those delivering services if immediate costs far outweigh immediate benefits. In global terms, staffing levels in US hospitals are high and so benefits from absolute staffing increases may be limited. Across all countries the economic arguments for increasing the proportion of Registered Nurses are more compelling than the argument for absolute increases and this applies in the US. A recent US study may shed some light on this. A panel study of over 2000 US hospitals found complex interactions and non-linear relationships between staffing level, outcomes, and costs. In simple terms increases in staffing were initially associated with reduced costs and improved outcomes. As staffing levels increased both associations were subject to tipping points so further increases in staffing became associated with increased costs and (at a higher level) no further improvements in outcomes.^45^

While there are limitations in the evidence, we judge that there is moderate certainty that our findings in relation to nurse staffing in general hospitals are correct. Despite weaknesses in the underlying evidence there is considerable degree of consistency in results. The potential cost-effectiveness of nurse staffing increases is in large part dependent on the magnitude of reduction in the risk of death for patients in acute hospitals. The effects across these economic studies, mostly based on cross-sectional associations, are consistent with those observed in longitudinal studies which are, in general, at much lower risk of bias.^4^ The most pervasive likely source of bias is simultaneity as staffing is likely to be increased in response to risk. This bias is likely to lead to an underestimate of the effect of staffing increases.^3 4^ We found very limited evidence about staffing in maternity settings and the cost-effectiveness of staffing increases cannot be determined. Although one US study found increased costs and no benefit, the context was very specific and both outcomes and costs considered were limited.

### 4.1 Limitations

The mortality-based outcomes considered for cost-effectiveness here are not the only value that can be delivered from increased staffing. While long term health gains might best be reflected in quality adjusted life gains these are insensitive measures and may not reflect important but less tangible benefits, for which individual healthcare consumers and society as a whole would still be willing to pay. Some of these benefits may be represented by improved patient experience and patient satisfaction, which are also associated with increased nurse staffing in several studies (e.g. ^42 46^). Specific conclusions about cost-effectiveness cannot be generalised, although a degree of consistency in results does give an indication of likely outcomes in other contexts.

Our searching was extensive, but the imprecise terminology and large number of potential studies means it is possible that some studies were missed. We were unable to assess publication bias but selective non reporting of results that are less favourable to nurse staffing is a possibility. However, it would require several studies with materially different results to change our conclusions.

### 4.2 Implications for policy & research

While there may be residual uncertainty around the cost-effectiveness of registered nurse staffing increases, the evidence of this review lends no support to policies that maintain or increase the size of the nursing workforce through skill mix dilution. In absolute terms the evidence is limited but the conclusions are clear. Increasing the proportion of registered nurses is associated with improved outcomes and reduced net cost. Conversely reducing skill mix increases costs and makes outcomes worse. Increased registered nurse staffing is potentially highly cost effective although local economic evaluations are required to establish incremental cost-effectiveness to inform decisions.

Studies of nurse staffing outcome associations continue to be published without any estimates of costs. The marginal utility of such studies is low and the priority for future research should be the use of more robust designs and the inclusion of economic evaluation using measures such as quality adjusted life years. As it seems likely that cost-effectiveness can be maximised by targeting staffing increases to areas of greatest need, more research is required to validate tools to guide such decisions, as current evidence is limited.^47^

### 4.3 Conclusions

In an era of registered nurse scarcity, our results strongly favour investment in registered nurse supply as opposed to using lesser qualified staff as substitutes. Our analysis gives strong support for increases in nurse skill mix and shows that policies that lead to a reduction in the proportion of registered nurses in nursing teams are likely to give worse outcomes at increased costs. Increases in absolute numbers of registered nurses in general medical wards have the potential to be highly cost effective, especially where baseline staffing is low, although more evidence on cost-effectiveness is still needed.

## Data Availability

All data produced in the present work are contained in the manuscript

## Acknowledgements & disclaimer

This study/project is funded by the NIHR [Health and Social Care Delivery Research (NIHR128056)] and the NIHR Applied Research Collaboration Wessex. The views expressed are those of the authors and not necessarily those of the NIHR or the Department of Health and Social Care.

