## Supplementary tables and appendix for "Costs and cost-effectiveness of nurse staffing in acute hospital: a systematic review"

### Supplementary Material

**Supplementary Table 7 Search strategy**

| **Database** | **Economics facet** | **Setting facet** | **Staffing facet** | **Staff groups facet** |
| --- | --- | --- | --- | --- |
| CINAHL | AB cost* OR  TI cost* OR  AB economic* OR  TI economic* OR  AB business case OR  TI business case | MH("Inpatients+") OR  MH ("Hospital Units+") OR  MH("Hospitals+") OR  AB Inpatient? OR  OR TI Inpatient? OR  AB Hospital? OR  TI Hospital? OR  AB(medical adj3 (unit* OR ward*)) OR  TI(medical adj3 (unit* OR ward*)) OR  AB(surgical adj3 (unit* OR ward*)) OR  TI(surgical adj3 (unit* OR ward*)) OR  AB(patient* adj3 surgical) OR  TI(patient* adj3 surgical) OR  AB(patient* adj3 medical) OR  TI(patient* adj3 medical) OR  AB("medical-surgical" OR "surgical-medical") OR  TI("medical-surgical" OR "surgical-medical") | (MH "Personnel Staffing and Scheduling+") OR  "staffing" OR  skill* N4 mix OR  staff* N3 (level* OR ratio* OR model* OR roster*) OR  "nursing hour* per patient day" OR  NHPPD OR  Workload OR  (MH "Workload") | (MH "Nurses+") OR  nurs* OR  (MH "Nursing Assistants") OR  (MH "RN First Assistants") OR  "nurs* aide*" OR  "healthcare assistant*" OR  "health care assistant*" OR  "healthcare support worker*" OR  "health care support worker*" OR  midwi* OR  (MH "Midwives+") OR  (MH "Midwifery Service+") |
| Embase through Ovid | Cost* OR  Economic* OR  Business Case).ab,ti. | Hospitals/ OR  exp Hospital Units/ OR  exp Inpatients/ OR  Inpatient?.ab,ti. OR  Hospital?.ab,ti. OR  (medical adj3 (unit* OR ward*)).ab,ti.OR  (surgical adj3 (unit* OR ward*)).ab,ti. OR  (patient* adj3 surgical).ab,ti. OR  (patient* adj3 medical).ab,ti. OR  ("medical-surgical" OR "surgical-medical").ab,ti. | staffing.mp. OR  exp Workforce/ OR  exp "Personnel Staffing and Scheduling"/ OR  (skill* adjModerate mix*).tw. OR  (staff* adj3 (level* or ratio* OR model* OR roster*)).mp.OR  exp Workload/ OR  workload.mp. | nurs*.mp. OR  Nursing Staff, Hospital/ OR  Nursing Services/ OR  exp Nursing Assistants/ OR  ("health care assistant*" OR "healthcare assistant*").mp.OR  ("health care support worker*" OR "healthcare support worker*").mp..OR  "midwi*".mp. OR  exp Midwifery/ |
| Econlit via EBSCO |  | AB Inpatient? OR  TI Inpatient? OR  AB Hospital? OR  TI Hospital? OR  AB(medical W3 (unit* OR ward*)) OR  TI(medical W3 (unit* OR ward*)) OR  AB(surgical W3 (unit* OR ward*)) OR  TI(surgical W3 (unit* OR ward*)) OR  AB(patient* W3 surgical) OR  TI(patient* W3 surgical) OR  AB(patient* W3 medical) OR  TI(patient* W3 medical) OR  AB("medical-surgical" OR "surgical-medical") OR  TI("medical-surgical" OR "surgical-medical") | "staffing" OR  skill* N4 mix OR  staff* N3 (level* OR ratio* OR model* OR roster*) OR  "nursing hour* per patient day" OR  NHPPD OR  Workload | nurs* OR  "nurs* aide*" OR  "healthcare assistant*" OR  "health care assistant*" OR  "healthcare support worker*" OR  "health care support worker*" OR  midwi* |
| Cochrane library: CDSR and CENTRAL and Cochrane Protocols | (cost*):ti,ab,kw OR  (economic*):ti,ab,kw OR  (business case):ti,ab,kw | MeSH descriptor: [Hospital units] explode all trees OR  MeSH descriptor: [Inpatients] explode all trees OR  MeSH descriptor: [Hospitals] explode all trees OR  (inpatient?):ti,ab,kw OR  (hospital?):ti,ab,kw OR  (medical near/3 (unit* OR ward*)):ti,ab,kw OR  (surgical near/3 (unit* OR ward*)):ti,ab,kw OR  (patient* near/3 surgical) :ti,ab,kw OR  (patient* near/3 medical) :ti,ab,kw OR  ("medical-surgical" OR "surgical-medical") :ti,ab,kw | MeSH descriptor: [Workforce] explode all trees OR  Staffing OR  MeSH descriptor: [Personnel Staffing and Scheduling] explode all trees | MeSH descriptor: [Nursing] explode all trees OR  nurs* OR  MeSH descriptor: [Midwifery] explode all trees OR  midwif* |
| DARE and NHS EED accessed through University of York CRD website | cost*:ti,ab OR  economic*:ti,ab OR  business case:ti,ab | MeSH descriptor: [Hospital units] explode all trees OR  MeSH descriptor: [Inpatients] explode all trees OR  MeSH descriptor: [Hospitals] explode all trees OR  (Inpatient?):ti,ab OR  (hospital?):ti,ab OR  (medical NEAR3 (unit* OR ward*)):ti,ab OR  (surgical NEAR3 (unit* OR ward*)):ti,ab OR  (patient* NEAR3 surgical) :ti,ab OR  (patient* NEAR3 medical) :ti,ab OR  ("medical-surgical" OR "surgical-medical") :ti,ab | MeSH descriptor: [Workforce] explode all trees OR  Staffing OR  MeSH descriptor: [Personnel Staffing and Scheduling] explode all trees | MeSH descriptor: [Nursing] explode all trees OR  nurs* OR  MeSH descriptor: [Midwifery] explode all trees OR  midwi* |
| HTA from INAHTA website | cost* [title,abstract,keywords] OR  economic* [title,abstract,keywords] OR  business case [title,abstract,keywords] | “Hospital units”[mhe] OR  “Inpatients”[mhe] OR  “Hospitals”[mhe] OR  Inpatient?[title,abstract,keywords] OR  Hospital?[title, abstract,keywords] OR  (medical W3 (unit* OR ward*))[title,abstract,keywords] OR  (surgical W3 (unit* OR ward*))[title,abstract,keywords] OR  (patient* W3 surgical)[title,abstract,keywords] OR  (patient* W3 medical) [title,abstract,keywords] OR  ("medical-surgical" OR "surgical-medical")[title,abstract,keywords] | “Workforce”[mhe] OR  Staffing OR  “Personnel Staffing and Scheduling”[mhe] OR  skill* W4 mix OR  staff* W3 (level* OR ratio* OR model* OR roster*) OR  (nursing hour* per patient day) OR  NHPPD OR  Workload | “Nursing”[mhe] OR  nurs* OR  “Midwifery” [mhe] OR  midwi* |
| PubMed | (Economic*[Title/Abstract]) OR  (Cost*[Title/Abstract]) OR  (Business case[Title/Abstract]) | (inpatients[MeSH Major Topic]) OR  (hospitals[MeSH Major Topic])) OR  (hospital units[MeSH Major Topic]) OR  inpatient?[Title/Abstract] OR  hospital?[Title/Abstract] OR  (medical-surgical[Title/Abstract]) OR (surgical-medical[Title/Abstract]) OR  "medical unit"[Title/Abstract] OR  "medical ward"[Title/Abstract] OR  "surgical unit"[Title/Abstract] OR  "surgical ward"[Title/Abstract] OR  "patient surgical"[Title/Abstract] OR  "patient medical"[Title/Abstract] | Staffing OR  "workforce"[MeSH Major Topic] OR  personnel staffing and scheduling"[MeSH Major Topic] OR  workload[MeSH Major Topic] OR  workload OR  "skill-mix"[Title/Abstract] OR  "skillmix"[Title/Abstract] OR  "skill-mix"[Title/Abstract] OR  ("staff*"[All Fields] AND "level*"[All Fields]) OR  ("staff*"[All Fields] AND "ratio*"[All Fields]) OR  ("staff*"[All Fields] AND "model*"[All Fields]) OR  ("staff*"[All Fields] AND "roster*"[All Fields]) | nurs* OR  "nursing staff, hospital"[MeSH Major Topic] OR  "nursing services"[MeSH Major Topic] OR  "nursing assistants"[MeSH Major Topic] OR  "health care assistant*"[All Fields] OR  "healthcare assistant*"[All Fields] OR  "health care support worker*"[All Fields] OR  "healthcare support worker*"[All Fields] OR  "midwi*"[All Fields] OR  "midwifery"[MeSH Major Topic] |

**Supplementary Table 6 Risk of bias**

| Study | Eligible population^[1]^ | Selected participant^[2]^s | **External validity*** | Design^[3]^ | Sufficient Power^[4]^ | Control for Confounding^[5]^ | **Internal validity*** | **Overall*** |
| --- | --- | --- | --- | --- | --- | --- | --- | --- |
| Behner et al. 1990 ^1^ | **-** | **++** | **-** | ++ | **-** | **-** | **-** | **-** |
| Clark et al. 2014 ^2^ | **+** | **++** | **+** | - | **++** | **-** | **-** | **-** |
| Cookson et al. 2014 ^3^ | **++** | **++** | **++** | - | **++** | **++** | **-** | **-** |
| Dall et al. 2009 ^4^ | **++** | **++** | **++** | - | **++** | **++** | **-** | **-** |
| Griffiths et al. 2018 ^5^ | **+** | **++** | **+** | ++ | **++** | **++** | **++** | **+** |
| Griffiths et al. 2020 ^6 7^ | **+** | **++** | **+** | ++ | **++** | **++** | **++** | **+** |
| Kim et al. 2016 ^8^ | **+** | **++** | **+** | - | **++** | **++** | **-** | **-** |
| Lasater et al. 2021a ^9^ | **+** | **+** | **+** | - | **++** | **++** | **-** | **-** |
| Lasater et al. 2021b ^10^ | **+** | **+** | **+** | + | **++** | **++** | **+** | **+** |
| Lasater et al. 2021c ^11^ | **+** | **+** | **+** | + | **++** | **++** | **+** | **+** |
| Li et al. 2011 ^12^ | **++** | **++** | **++** | **-** | **++** | **++** | **-** | **-** |
| Li et al. 2016 ^13^ | **+** | **++** | **+** | + | **++** | **++** | **+** | **+** |
| Martsolf et al. 2014 ^14^ | **++** | **++** | **++** | - | **++** | **+** | **-** | **-** |
| Needleman et al. 2006 ^15^ | **++** | **++** | **++** | - | **++** | **++** | **-** | **-** |
| Pang et al. 2019 ^16^ | **+** | **++** | **+** | - | **+** | **+** | **-** | **-** |
| Ross et al. 2021 ^17^ | **+** | **++** | **+** | - | **++** | **++** | **-** | **-** |
| Rothberg et al. 2005 ^18^ (c) | **+** | **+** | **+** | - | **++** | **+** | **-** | **-** |
| Shamliyan et al. 2009 ^19^ (d) | **++** | **++** | **++** | + | **++** | **+** | **+** | **+** |
| Twigg et al. 2013 ^20^ | **+** | **++** | **+** | + | **++** | **++** | **+** | **+** |
| Van den Heede et al. 2010 ^21^ | **++** | **++** | **++** | - | **+** | **++** | **-** | **-** |
| Weiss et al. 2011 ^22^ | **+** | **+** | **+** | - | **+** | **++** | **-** | **-** |
| Yakusheva et al. 2014 ^23^ | **+** | **++** | **+** | ++ | **+** | **++** | **+** | **+** |

- *Summary rating based on lowest score in preceding section-
- - high risk of bias (weak), + moderate (stronger), ++ low risk of bias (strong)

Appendix:

**Operationalisation of risk of bias assessment**

Guidance on risk of bias assessment for studies of association – based on items from a framework for the development of NICE public health guidance as adapted for studies of staffing association. ^24 25^

*Items rated & guidance on application & interpretation:*

**Primarily related to external validity (can the estimates from the study be generalised)**

- **Is the eligible population or area representative of the source population or area?**

Interpretation: The source population is interpreted as all inpatients cared for by staff included in the staffing variable (and hence to whom inferences about benefits and costs might be made).

Where the sample is of patients representative of the whole service (+) and from multiple hospitals (≥Moderate0) representative of a health system (+) (national or regional) rate as strong (++). If there is a mismatch between the patient group and staff (e.g. associations between hospital staffing and outcomes from a patient sub-group) rate down (-) as the target population for the study does not represent the target population for the service.

Studies in Moderate0 or more hospitals where a relatively small subgroup of patients would (only) score ++ if the staffing data is from units occupied by those patients AND those patients are representative of all patients in those units. Where staffing outcomes relationships are derived from systematic reviews assume ++ if most underlying studies in the review meet the criteria.

- **Do the selected participants or areas represent the eligible population or area?**

Interpretation: Consider whether staffing data is complete and whether all eligible patients are included. Where staff (+) and patient (+) data is derived from administrative systems rate as strong (++). If staffing or patient data is based on other data collections (e.g. a survey or audit) rate as + only if there is good evidence of near complete (80%) response from the eligible populations such that there is assurance that for each unit included in the study both patient and staff responses are 80%+.

Summary risk of bias based on lowest scoring aspect

**Primarily related to internal validity (are the inferences from the study likely to be correct)**

- **Strength of research design**

Interpretation: Cross-sectional studies where associations between patient outcomes and staffing aggregated over time / across a whole facility to be rated as weak (-) unless there is evidence of matching or comparable approaches to ensure comparability of the exposed samples (+). Where staffing and patient data are linked at the unit / day level (or similar) in a longitudinal fashion rate as strong (++). Where staffing outcomes relationships are derived from systematic reviews rate as strong (++) if the evidence includes longitudinal studies and the review has considered design sensitivity.

- **Sufficient Power**

Interpretation: As studies rarely provided a power calculation and effect sizes are anticipated to be small, but small effects may be clinically important (especially at a service level) large samples are required. In order to give an approximate relative indication of power studies with <Moderate000 patients were rated as weak <Moderate000 and those with Moderate0,000 or more as strong.

- **How well were likely confounding factors identified and controlled?**

Interpretation: Studies that included a clearly defined risk adjustment approach for the main outcome including key factors such as age, co-morbidities and diagnostic information or use an otherwise validated risk adjustment model were rated as strong (++). Studies without patient level risk adjustment were rated 0 or + if there was evidence of stratification of units that accounted for patient risk or unit level case mix factors were included.

Summary risk of bias based on lowest scoring aspect

**Studies excluded at full text review stage (with reasons)**

| Author [citation] |  | Title |  | Reason excluded |
| --- | --- | --- | --- | --- |
| New South Wales Nurses' Association [1] |  | Ratios must be at heart of health debate |  | Discussion / review |
| Adams & Kaplow [2] |  | A Sitter-Reduction Program In an Acute Health Care System |  | Not staffing – outcomes / cost |
| Allen et al.[3] |  | The impact of caseload midwifery, compared with standard care, on women's perceptions of antenatal care quality: survey results from the M@NGO randomized controlled trial for women of any risk | | Staffing model |
| Alorbi [4] |  | Essays on Hospital Reimbursement and Quality of Healthcare Provision |  | Not staffing – outcomes / cost |
| Altman et al. [5] |  | The Cost of Nurse-Midwifery Care: Use of Interventions, Resources, and Associated Costs in the Hospital Setting |  | Staffing model |
| Ball et al. [6] |  | Birthrate Plus Programme. Factors affecting staffing ratios |  | Staffing model |
| Bateman et al. [7] |  | 36th International Symposium on Intensive Care and Emergency Medicine |  | Abstract (no data) |
| Bellanger and Or [8] |  | What can we learn from a cross-country comparison of the costs of child delivery? |  | Staffing model |
| Bhattacharyya et al. [9] |  | A unique orthogeriatric model: a step forward in improving the quality of care for hip fracture patients |  | Not staffing - outcomes |
| Birnbaum et al. [10] |  | Facilitating a Major Staffing Transition in a State Psychiatric Hospital With Changes to Nursing Orientation |  | Ineligible service |
| Bonafide et al. [11] |  | Cost-Benefit Analysis of a Medical Emergency Team in a Children’s Hospital |  | Can't isolate RN |
| Bowers et al. [12] |  | A multicriteria resource allocation model for the redesign of services following birth |  | Ineligible service |
| Brockman [13] |  | Implementing the Mother-Baby Model of Nursing Care Using Models and Quality Improvement Tools |  | Staffing model |
| Butler et al. [14] |  | Hospital nurse‐staffing models and patient‐ and staff‐related outcomes |  | Discussion / review |
| Carter [15] |  | Nursing Care Left Undone in European Hospitals |  | Discussion / review |
| Chang et al. [16] |  | The Relationship Between Nursing Workload, Quality of Care, and Nursing Payment in Intensive Care Units |  | Not staffing – outcomes /costs |
| Cho et al. [17] |  | The effects of nurse staffing on adverse events, morbidity, mortality, and medical costs |  | Not staffing – outcomes /costs |
| Collins et al. [18] |  | Outcomes of adding acute care nurse practitioners to a Level I trauma service with the goal of decreased length of stay and improved physician and nursing satisfaction | | Staffing model |
| Delhy et al. [19] |  | The Impact of Nursing Staff on Satisfaction Scores for U.S. Hospitals: A Production Function Approach |  | No economic data |
| DeRienzo et al. [20] |  | A discrete event simulation tool to support and predict hospital and clinic staffing |  | No economic data |
| Drake [21] |  | Nursing workforce planning: insights from seven Malaysian hospitals |  | No economic data |
| Dzikowicz et al. [22] |  | Comparing an All-RN Unit to a Mixed-Skill Unit at a Hospital |  | No economic data |
| Edkins et al. [23] |  | A systematic review of advance practice providers in acute care: options for a new model in a burn intensive care unit |  | Discussion / review |
| Ferdosi et al. [24] |  | Evaluation of outsourcing in nursing services: a case study of kashani hospital, isfahan in 2011 |  | Not staffing – outcomes /costs |
| Freeman et al. [25] |  | Gatekeepers at Work: An Empirical Analysis of a Maternity Unit |  | No economic data |
| Guido-Sanz [26] |  | The effects of advanced practice nurses (APNs) as intensivists in a surgical intensive care unit (SICU) on patient outcomes, healthcare charges, and apn intensivist services in the SICU | | Staffing model |
| Hairr et al. [27] |  | Nurse Staffing and the Relationship to Job Satisfaction And Retention |  | No economic data |
| Jacob et al. [28] |  | The changing skill mix in nursing: considerations for and against different levels of nurse |  | Discussion / review |
| Kapu et al [29] |  | FInancial impact of adding nurse practitioners (NPS) to critical care teams |  | Abstract (no data) |
| Kapu and Steaban [30] |  | Adding Nurse Practitioners to Inpatient Teams: Making the Financial Case for Success |  | No economic data |
| Kartha and McCrone [31] |  | Cost-effectiveness of staffing levels on conflict and containment on psychiatric wards in England |  | Ineligibble service |
| Kokangul et al. [32] |  | Optimizing Nurse Capacity in a Teaching Hospital Neonatal Intensive Care Unit |  | No economic data |
| Liang et al. [33] |  | Estimating a Hospital Production Function to Evaluate the Effect of Nurse Staffing on Patient Mortality in Taiwan: The Longitudinal Count Data Approach | | No economic data |
| Min [34] |  | Evaluation of Technical Efficiency of Nursing Care in U.S. Critical Care Units |  | No economic data |
| Needleman [35] |  | The Economic Case for Fundamental Nursing Care |  | Discussion / review |
| Newbold [36] |  | The production economics of nursing: a discussion paper |  | Discussion / review |
| Pappas et al. [37] |  | Risk-Adjusted Staffing to Improve Patient Value |  | No economic data |
| Roberts et al. [38] |  | Use of health economic evaluation in the implementation and improvement science fields-a systematic literature review |  | Discussion / review |
| Sandall et al. [39] |  | The efficient use of the maternity workforce and the implications for safety and quality in maternity care: a population-based, cross-sectional study | | No economic data |
| Serratt [40] |  | California's nurse-to-patient ratios, Part 2: 8 years later, What do we know about hospital level outcomes? |  | Discussion / review |
| Togeiro Fugulin et al. [41] |  | Nursing staff in the internal medicine and surgical units of teaching hospitals: composition and cost |  | No economic data |
| Tracy et al. [42] |  | Caseload midwifery compared to standard or private obstetric care for first time mothers in a public teaching hospital in Australia: A cross sectional study of cost and birth outcomes | | Staffing model |
| Twigg et al. [43] |  | Is there an economic case for investing in nursing care--what does the literature tell us? |  | Discussion / review |
| Watson et al. [44] |  | The effects of a one-to-one nurse-to-patient ratio on the mortality rate in neonatal intensive care: A retrospective, longitudinal, population-based study | | No economic data |
| Yoder [45] |  | AMSN President's Message. Making the Business Case for Medical-Surgical Nurses |  | Discussion / review |

**Full references**

1. *Ratios must be at heart of health debate.* Lamp, 2015. **72**(2): p. 20-21.
2. Adams, J. and R. Kaplow, *A Sitter-Reduction Program In an Acute Health Care System.* Nursing Economic$, 2013. **31**(2): p. 83-89.
3. Adams, W., *Standards and recommendations that enhance the quality of ambulatory surgery.* Ambulatory Surgery, 2015. **21 (2)**: p. 34.
4. Alorbi, G.A., *Essays on Hospital Reimbursement and Quality of Healthcare Provision*. 2017, Southern Illinois University.
5. Altman, M.R., et al., *The Cost of Nurse-Midwifery Care: Use of Interventions, Resources, and Associated Costs in the Hospital Setting.* Womens Health Issues, 2017. **27**(4): p. 434-440.
6. Ball, J., et al., *Birthrate Plus Programme. Factors affecting staffing ratios.* British Journal of Midwifery, 2003. **11**(6): p. 357-360.
7. Bateman, R.M., et al., *36th International Symposium on Intensive Care and Emergency Medicine : Brussels, Belgium. 15-18 March 2016.* Crit Care, 2016. **20**(Suppl 2): p. 94.
8. Bellanger, M.M. and Z. Or, *What can we learn from a cross‐country comparison of the costs of child delivery?* Health economics, 2008. **17**(S1): p. S47-S57.
9. Bhattacharyya, R., et al., *A unique orthogeriatric model: a step forward in improving the quality of care for hip fracture patients.* Int J Surg, 2013. **11**(10): p. 1083-6.
10. Birnbaum, S., et al., *Facilitating a Major Staffing Transition in a State Psychiatric Hospital With Changes to Nursing Orientation.* Journal of the American Psychiatric Nurses Association, 2017. **23**(1): p. 66-72.
11. Bonafide, C.P., et al., *Cost-Benefit Analysis of a Medical Emergency Team in a Children’s Hospital.* Pediatrics, 2014. **134**(2): p. 235-241.
12. Bowers, J., et al., *A multicriteria resource allocation model for the redesign of services following birth.* BMC Health Serv Res, 2018. **18**(1): p. 656.
13. Brockman, V., *Implementing the Mother-Baby Model of Nursing Care Using Models and Quality Improvement Tools.* Nurs Womens Health, 2015. **19**(6): p. 490-503.
14. Butler, M., et al., *Hospital nurse‐staffing models and patient‐ and staff‐related outcomes.* Cochrane Database of Systematic Reviews, 2019(4).
15. Carter, D., *Nursing Care Left Undone in European Hospitals.* AJN American Journal of Nursing, 2014. **114**(2): p. 17-17.
16. Chang, L.Y., H.H. Yu, and Y.F.C. Chao, *The Relationship Between Nursing Workload, Quality of Care, and Nursing Payment in Intensive Care Units.* The journal of nursing research : JNR, 2019. **27**(1): p. 1-9.
17. Cho, S., et al., *The effects of nurse staffing on adverse events, morbidity, mortality, and medical costs.* Nursing Research, 2003. **52**(2): p. 71-79.
18. Collins, N., et al., *Outcomes of adding acute care nurse practitioners to a Level I trauma service with the goal of decreased length of stay and improved physician and nursing satisfaction.* J Trauma Acute Care Surg, 2014. **76**(2): p. 353-7.
19. Delhy, R., A. Dor, and P. Pittman, *The Impact of Nursing Staff on Satisfaction Scores for U.S. Hospitals: A Production Function Approach.* Medical Care Research and Review., 2020.
20. DeRienzo, C.M., et al., *A discrete event simulation tool to support and predict hospital and clinic staffing.* Health Informatics Journal, 2017. **23**(2): p. 124-133.
21. Drake, R., *Nursing workforce planning: insights from seven Malaysian hospitals.* British Journal of Nursing, 2013. **22**(2): p. 95-100.
22. Dzikowicz, D.J., et al., *Comparing an All-RN Unit to a Mixed-Skill Unit at a Hospital.* The Journal of nursing administration, 2020. **50**(12): p. e14-e22.
23. Edkins, R.E., B.A. Cairns, and C.S. Hultman, *A systematic review of advance practice providers in acute care: options for a new model in a burn intensive care unit.* Annals of plastic surgery, 2014. **72**(3): p. 285-288.
24. Ferdosi, M., et al., *Evaluation of outsourcing in nursing services: a case study of kashani hospital, isfahan in 2011.* Mater Sociomed, 2013. **25**(1): p. 37-9.
25. Freeman, M., N. Savva, and S. Scholtes, *Gatekeepers at Work: An Empirical Analysis of a Maternity Unit.* Management Science, 2017. **63**(10): p. 3147-3167.
26. Guido-Sanz, F., *The effects of advanced practice nurses (APNs) as intensivists in a surgical intensive care unit (SICU) on patient outcomes, healthcare charges, and apn intensivist services in the SICU*. 2014, Florida International University. p. 221 p-221 p.
27. Hairr, D.C., et al., *Nurse Staffing and the Relationship to Job Satisfaction And Retention.* Nursing Economic$, 2014. **32**(3): p. 142-147.
28. Jacob, E.R., L. McKenna, and A. D'Amore, *The changing skill mix in nursing: considerations for and against different levels of nurse.* Journal of Nursing Management (John Wiley & Sons, Inc.), 2015. **23**(4): p. 421-426.
29. Kapu AN, Steaban R. Adding nurse practitioners to inpatient teams: Making the financial case for success. Nurse Leader. 2016;14(3):198-202.
30. Kapu, A. and P. Jones, *Financial impact of adding acute care nurse practitioners (ACNPs) to inpatient models of care.* Critical Care Medicine, 2012. **40**(12): p. 27.
31. Kartha, M.R. and P. McCrone, *Cost-effectiveness of staffing levels on conflict and containment on psychiatric wards in England.* Journal of psychiatric and mental health nursing, 2019. **26**(9-10): p. 337-346.
32. Kokangul, A., S. Akcan, and M. Narli, *Optimizing Nurse Capacity in a Teaching Hospital Neonatal Intensive Care Unit.* Health Care Management Science, 2017. **20**(2): p. 276-285.
33. Liang, Y.-W., W.-Y. Chen, and Y.-H. Lin, *Estimating a Hospital Production Function to Evaluate the Effect of Nurse Staffing on Patient Mortality in Taiwan: The Longitudinal Count Data Approach.* Romanian Journal of Economic Forecasting, 2015. **18**(4): p. 154-169.
34. Min, A., *Evaluation of Technical Efficiency of Nursing Care in U.S. Critical Care Units*, in *Evaluation of Technical Efficiency of Nursing Care in U.s. Critical Care Units*. 2016. p. 1-1.
35. Needleman, J., *The Economic Case for Fundamental Nursing Care.* Nursing Leadership (1910-622X), 2016. **29**(1): p. 26-36.
36. Newbold, D., *The production economics of nursing: a discussion paper.* Int J Nurs Stud, 2008. **45**(1): p. 120-8.
37. Pappas, S., et al., *Risk-Adjusted Staffing to Improve Patient Value.* Nursing economic$, 2015. **33**(2): p. 73-8, 87; quiz 79.
38. Roberts, S.L.E., A. Healey, and N. Sevdalis, *Use of health economic evaluation in the implementation and improvement science fields-a systematic literature review.* Implementation Science, 2019. **14**(1): p. N.PAG-N.PAG.
39. Sandall, J., et al., *The efficient use of the maternity workforce and the implications for safety and quality in maternity care: a population-based, cross-sectional study* Health Services and Delivery Research, 2014. **2**(38): p. 10.3310/hsdr02380.
40. Serratt, T., *California's nurse-to-patient ratios, Part 2: 8 years later, What do we know about hospital level outcomes?* Journal of Nursing Administration, 2013. **43**(10): p. 549-553.
41. Togeiro Fugulin, F.M., et al., *Nursing staff in the internal medicine and surgical units of teaching hospitals: composition and cost.* Revista da Escola de Enfermagem da USP, 2015. **49**: p. 47-53.
42. Tracy, S.K., et al., *Caseload midwifery compared to standard or private obstetric care for first time mothers in a public teaching hospital in Australia: A cross sectional study of cost and birth outcomes.* BMC Pregnancy and Childbirth, 2014. **14**(1).
43. Twigg, D.E., et al., *Is there an economic case for investing in nursing care--what does the literature tell us?* Journal of advanced nursing, 2015. **71**(5): p. 975-990.
44. Watson, S.I., et al., *The effects of a one-to-one nurse-to-patient ratio on the mortality rate in neonatal intensive care: A retrospective, longitudinal, population-based study.* Archives of Disease in Childhood: Fetal and Neonatal Edition, 2016. **101**(3): p. F195-F200.
45. Yoder, L., *AMSN President's Message. Making the Business Case for Medical-Surgical Nurses.* MEDSURG Nursing, 2018. **27**(2): p. 73-74.

24. National Institute for Clinical Excellence. Methods for the development of NICE public health guidance: NICE: London 2012.

25. Griffiths P, Ball J, Drennan J, et al. The association between patient safety outcomes and nurse/healthcare assistant skill mix and staffing levels and factors that may influence staffing requirements (NICE evidence review): University of Southampton Centre for innovation and Leadership in Health Sciences, 2014.

1. Target population is patients in service for which staffing is measured. If the patient group included represents the target population and the sample of hospitals is 10 or more rate as strong.
2. If staffing data is complete and the target patient population is fully sampled (e.g. administrative data) rate as strong
3. Cross-sectional designs rated as weak unless accompanied by additional features such as matching / propensity (stronger). Longitudinal studies and studies with using individual patient exposure measures rated as strong
4. Based on sample size alone as no power calcs given - small (weak) <1000< medium (stronger)<10000 large (strong) 10,000+ patients
5. No adjustment for patient risk factors (weak) partial (stronger) or extensive (strong)
